# Genome-wide Screen of Otosclerosis in Population Biobanks: 18 Loci and Shared Heritability with Skeletal Structure

**DOI:** 10.1101/2020.11.15.20227868

**Authors:** Joel T. Rämö, Tuomo Kiiskinen, Juha Karjalainen, Kristi Krebs, Mitja Kurki, Aki S. Havulinna, Eija Hämäläinen, Paavo Häppölä, Heidi Hautakangas, FinnGen, Konrad J. Karczewski, Masahiro Kanai, Reedik Mägi, Priit Palta, Tõnu Esko, Andres Metspalu, Matti Pirinen, Samuli Ripatti, Lili Milani, Antti Mäkitie, Mark J. Daly, Aarno Palotie

## Abstract

Otosclerosis is one of the most common causes of conductive hearing loss, affecting 0.3% of the population. It typically presents in adulthood and half of the patients have a positive family history. The pathophysiology of otosclerosis is poorly understood and treatment options are limited. A previous genome-wide association study (GWAS) identified a single association locus in an intronic region of *RELN*. Here, we report a meta-analysis of GWAS studies of otosclerosis in three population-based biobanks comprising 2,413 cases and 762,382 controls. We identify 15 novel risk loci (p < 5*10^−8^) and replicate the regions of *RELN* and two previously reported candidate genes (*TGFB1* and *MEPE*). Implicated genes in many loci are essential for bone remodelling or mineralization. Otosclerosis is genetically correlated with height and fracture risk, and the association loci overlap with severe skeletal disorders. Our results highlight TGFβ1 signalling for follow-up mechanistic studies.

## Introduction

Otosclerosis is one of the most common causes of conductive hearing loss. It is a heritable disorder typically characterized by disordered bone growth in the middle ear.^1,2^ In classic otosclerosis, due to fixation of the footplate of the stapes bone, the conduction of sound through the ossicular chain to the inner ear is interrupted. The pathogenesis of otosclerosis is poorly understood, and therapeutic options are mostly limited to hearing aids, prosthesis surgery and cochlear implants.^3^

Clinical otosclerosis has an estimated prevalence of 0.30% to 0.38% in Caucasian populations.^4^ Histologic otosclerosis without clinical symptoms is more frequent, with bony overgrowth observed in as many as 2.5% of temporal bone autopsy specimens.^5^ Symptomatic otosclerosis most frequently occurs in working-age individuals between the second and fifth decades.^6,7^ Initial manifestation is often limited to one ear, but eventual bilateral disease is observed in 70-80% of cases.^6,7^

Otosclerosis is highly familial, with a positive family history reported for 50-60% of cases.^8,9^ Based on segregation patterns, early studies classified otosclerosis as an autosomal dominant disease with reduced penetrance.^9-11^ However, efforts to identify causative genes have produced inconsistent results, with insufficient evidence for most candidate genes.^12,13^ A genome-wide association study (GWAS) including a total of 1,149 otosclerosis patients identified intronic risk variants within the gene encoding Reelin (*RELN)* in European populations.^14,15^ However, the potential biological function of Reelin and the role of other genes remain unknown.

Here, we report to our knowledge the largest GWAS of otosclerosis including a total of 2,413 cases and 762,382 controls from three population-based sample collections: the Finnish FinnGen study, the Estonian Biobank (EstBB), and the UK Biobank (UKBB). We identify 15 novel GWAS loci, and replicate associations in the regions of *RELN* and two previously reported candidate genes, *TGFB1* and *MEPE*.^14-19^ Several discovered loci harbor genes involved in the regulation of osteoblast or osteoclast function or biomineralization. We also demonstrate that otosclerosis is genetically correlated with common and rare skeletal traits.

## Results

### Identification of otosclerosis cases based on electronic health records

Based on International Clasification of Diseases (ICD) diagnosis codes (versions 8, 9 and 10), we identified a total of 2,413 otosclerosis cases in the three biobanks, including 1,350 cases in FinnGen, 713 in EstBB and 353 in UKBB, for cohort prevalences of 0.64%, 0.52%, and 0.08%, respectively (Table 1). A total of 762,382 individuals had no ICD-based diagnosis of otosclerosis and were assigned control status. A proportion of otosclerosis patients undergo stapes procedures, which reflect disease severity and the validity of the ICD-based diagnoses. Stapes procedures were registered for 544 (40.3%) otosclerosis cases in FinnGen and for 161 (22.6%) otosclerosis cases in EstBB. Among all 1,350 individuals diagnosed with otosclerosis in FinnGen, five (0.37%) had a diagnosis of osteogenesis imperfecta (representing 12% of all 41 individuals with osteogenesis imperfecta), and none had a diagnosis of osteopetrosis.

**Table 1.** ICD and procedure codes corresponding to otosclerosis in each study cohort. Stapes procedures were identified based on the Nomesco codes DDA00 (stapedotomy) and DDB00 (stapedectomy) in FinnGen and the national health insurance treatment service code 61006 (stapedotomy) in EstBB.

| Classification | Code | Definition | FinnGen | EstBB | UKBB |
| --- | --- | --- | --- | --- | --- |
| ICD-10 | H80 | Otosclerosis (any) |  | 256 | 318 |
|  | H80.0 | .. Involving oval window, nonobliterative | 655 | 246 | 10 |
|  | H80.1 | .. Involving oval window, obliterative | 185 | 63 | 3 |
|  | H80.2 | .. Cochlear | 45 | 32 | 3 |
|  | H80.8 | .. Other | 133 | 95 | 6 |
|  | H80.9 | .. Unspecified | 628 | 379 | 301 |
| ICD-9 | 387 | Otosclerosis (any) | 272 |  | 36 |
| ICD-8 | 386 | Otosclerosis (any) | 284 |  |  |
| Procedures |  | Stapedotomy | 458 | 161 |  |
| Procedures |  | Stapedectomy | 113 |  |  |
| **Total cases** |  |  | **1350** | **713** | **350** |
| **Total controls** |  |  | **209582** | **136793** | **416007** |
| Prevalence (%) |  |  | 0.64 | 0.52 | 0.08 |

### Genomic loci associated with otosclerosis in each cohort

In case-control GWASs within the individual cohorts, we observed ten loci associated with otosclerosis at genome-wide significance (*p* < 5*10^−8^) in FinnGen, four in UKBB and one in EstBB (Supplementary Data 1; Supplementary Data 2). Two loci were associated in both FinnGen and UKBB: A chromosome 11 locus tagged by the lead variants rs11601767 and rs144458122, respectively, and a chromosome 17 locus upstream of *FAM20A* tagged by rs11868207 and rs11652821. Among all FinnGen lead variants, six were associated with otosclerosis at nominal significance (*p* < 0.05) in UKBB and five were associated with otosclerosis in EstBB. All three UKBB lead variants were nominally significant in FinnGen. The only genome-wide significant variant in EstBB (rs78942990, *p* = 4.2 * 10^−8^, MAF = 2.0%) was not associated with otosclerosis in either FinnGen or UKBB at nominal significance level.

We calculated pairwise genetic correlation r_g_ using summary statistics from the different cohorts with the cross-trait LD Score Regression.^20,21^ R_g_ between FinnGen and UKBB was 1.04, indicating similar genetic effects between the two cohorts.^22^ The calculation of r_g_ was not feasible for EstBB due to a negative heritability estimate (observed scale h^2^ = -0.0063 [S.E. 0.003]), likely reflecting modest polygenic signal in the cohort. However, as the effect estimates for lead variants were largely concordant between the three cohorts (Supplementary Data 2), we proceeded with a fixed-effects meta-analysis.

### 18 significant loci in the meta-analysis of summary statistics

In the meta-analysis of all three cohorts (including a total of 2,413 cases and 762,382 controls) we identified 1,257 variants associated with otosclerosis at the genome-wide significance level (p < 5*10^−8^). The genomic inflation factor calculated from the meta-analysis *p*-values was not significantly elevated (λ_GC_=1.03). The univariate LD Score regression intercept was closer to 1.0 at 1.016 (S.E. 0.009), indicating that part of the elevation in λ_GC_ reflected true additive polygenic effects and not spurious associations from population stratification or cryptic relatedness.^21^

The significant variants in the meta-analysis clustered in a total of 18 loci (regions at least 500kb apart) (Figure 1; Table 2; Supplementary Data 3; Supplementary Data 4), including the previously reported *RELN* locus and 17 loci not previously reported in a GWAS study of otosclerosis.^14^ All lead variants in the 18 meta-analysis loci were common with a cross-cohort effect allele frequency (EAF) of at least 16% (Table 2). In two loci the closest genes (*MEPE* and *TGFB1*) have been implicated in candidate gene studies; to our knowledge, the remaining 15 loci have not been characterized in association with otosclerosis.^15-19^

**Table 2.**
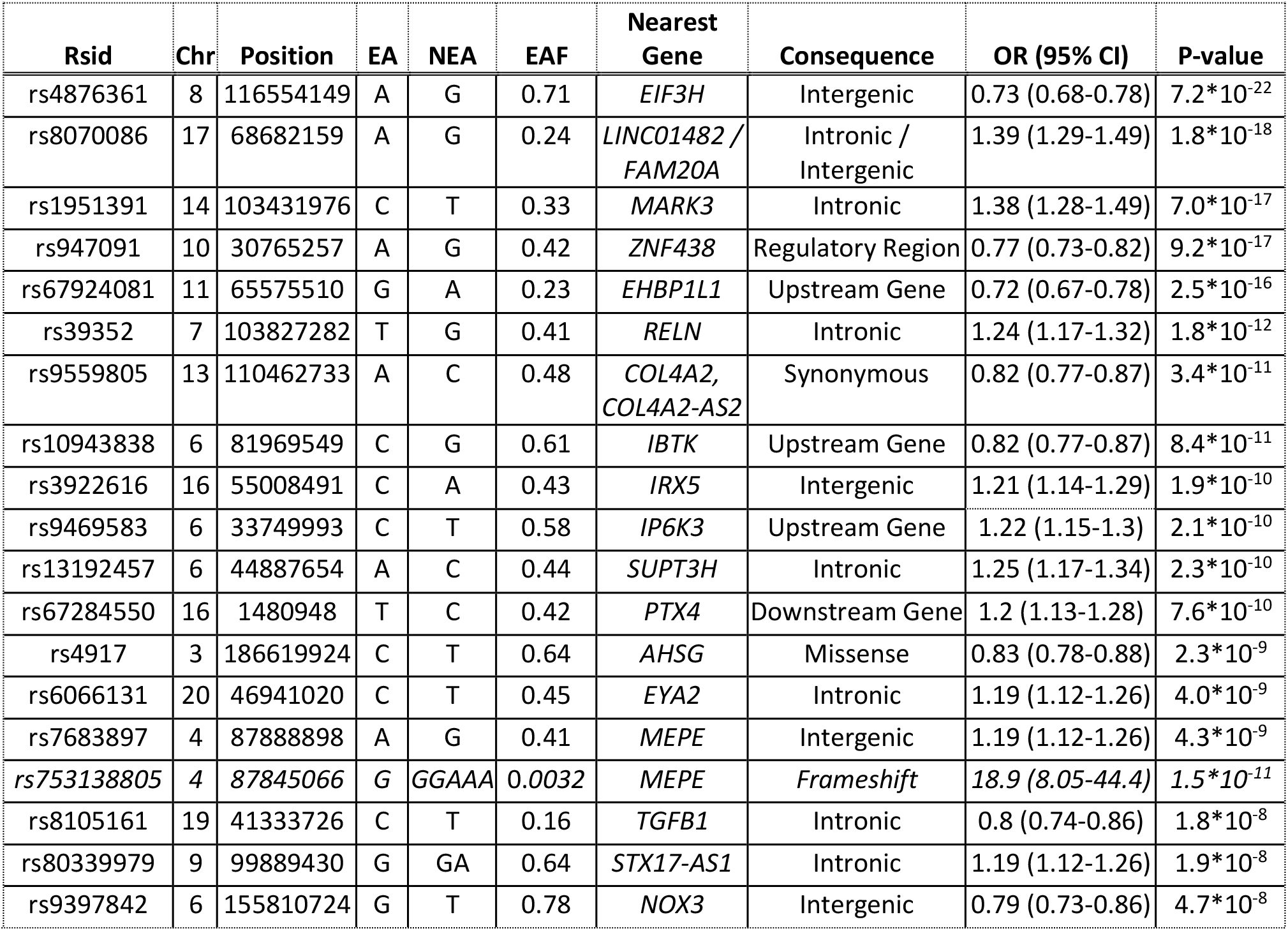
Lead variants for significant association loci in the meta-analysis of otosclerosis. Data (in cursive) are also presented for the rs753138805 variant in *MEPE*, observed only in FinnGen and not included in the meta-analysis. *Chr = chromosome, EA = effect allele, EAF = effect allele frequency, NEA = non-effect allele, OR = odds ratio, CI = confidence interval*.

**Figure 1.**
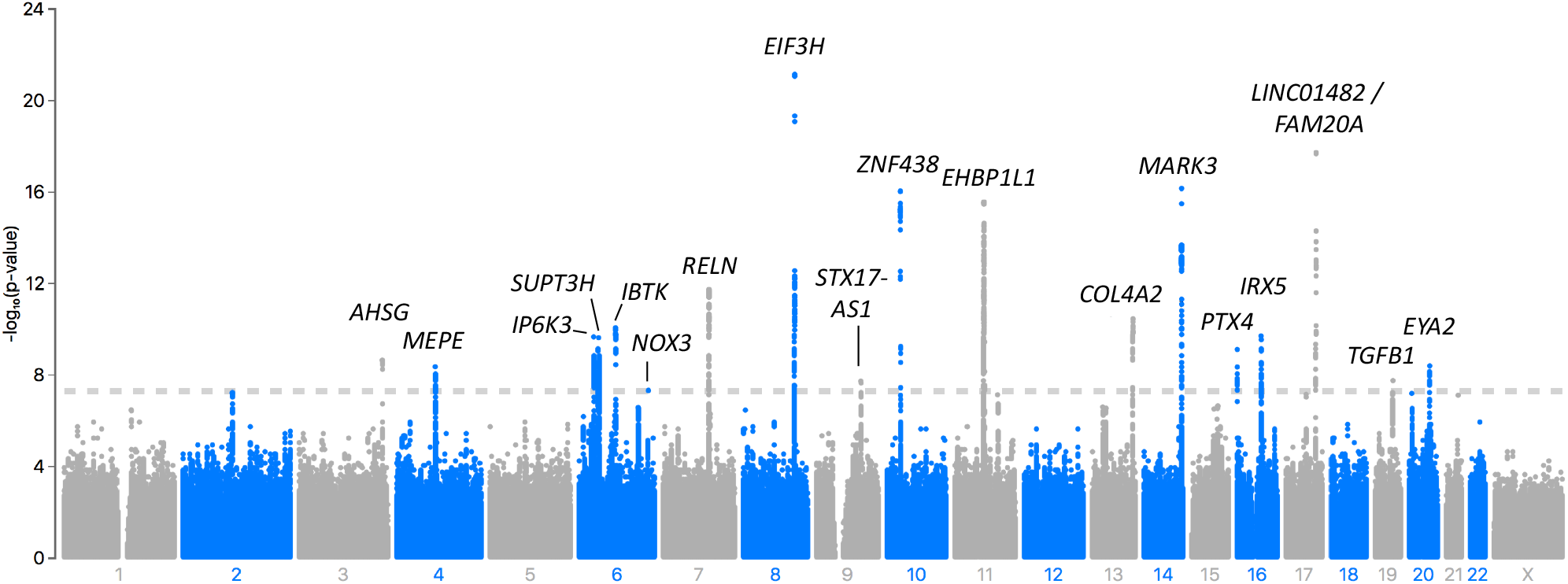
Meta-analysis of the genome-wide association studies of Otosclerosis in FinnGen, EstBB and UKBB. In the Manhattan plot, chromosomal positions are indicated on the x-axis and -log_10_(p-value) is presented on the y-axis for each variant. Results are presented for a fixed-effect meta-analysis of effect estimates from GWASs in the three cohorts, including a total of 2,413 cases and 762,382 controls. The included variants were present in at least two cohorts with a cross-cohort minor allele frequency > 0.1% and imputation INFO score > 0.7. Eighteen loci reached genome-wide significance (p < 5*10^−8^, marked by the dashed line). The loci are annotated by the names of the genes nearest to the lead variants.

### Characterization of the potential susceptibility loci for otosclerosis

#### Coding variation

The lead variant in the chromosome 3 locus, rs4917 T>C, is a missense variant in the exon 6 of *AHSG* (EAF = 64%, OR = 0.83 [0.78-0.88], *p* = 2.3*10^−9^). The variant results in a methionine to threonine conversion and is predicted to be tolerated/benign by the SIFT and PolyPhen algorithms with a scaled CADD score of 9.9. However, it has been reported as a splicing quantitative trait locus (sQTL) for AHSG in the liver in the GTEx v8 database (*p* = 1.1 * 10^−7^).^23^ Among all 1,257 variants which reached genome-wide significance in the meta-analysis, we identified no high-impact variants. Seven missense variants were significantly associated with otosclerosis (Supplementary Data 5): rs1193851 C>G in *PCNX3*, rs11544832 in *YIF1A*, rs7990383 G>A in *COL4A2*, rs7990383 C>A in *IRX5*, rs4918 G>C in *AHSG*, rs2229642 C>G in *ITPR3*, and rs4713668 in *IP6K3*. Rs4713668 (EAF = 0.48, OR = 1.19 [1.12-1.26], *p* = 8.8*10^−9^) is classified as deleterious and benign by the SIFT and PolyPhen algorithms, respectively, with a scaled CADD score of 19.9. All other missense variants are classified as tolerated or benign.

In the chromosome 4 locus near the *MEPE* gene, the rare frameshift variant rs753138805 (EAF 0.3% in controls and 1.3% in cases) was the lead variant in the GWAS in FinnGen, but was not included in the meta-analysis due to poor imputation quality in EstBB (Imputation information score 0.28) and non-inclusion in UKBB genotype data. The rs753138805 variant was strongly associated with otosclerosis in FinnGen (OR = 18.9 [95% CI 8.05-44.4]; *p* = 1.5*10^−11^). The variant is 2.4-fold enriched in Finnish compared with non-Finnish Europeans based on sequence data in the Genome Aggregation Database.^24^ We validated the imputation of the variant within the Migraine Family subcohort of FinnGen. Among 65 individuals determined as heterozygous for rs753138805 by imputation, we confirmed the genotype by Sanger sequencing in 56 individuals (86%) and in all five predicted carriers of the reference allele examined as controls.

Finally, based on linkage disequilibrium in the FinnGen cohort, we examined 988 variants (including those with AF < 0.001) in high LD (r > 0.6) with the lead variants from the meta-analysis, but observed no additional protein-altering variation.

#### Intronic and intergenic variation

The lead variants were intronic in seven loci. In the locus on chromosome 7, the strongest association was observed for variants in the second and third introns of *RELN* concordant with previous GWA studies (Supplementary Data 4).^14,15^ In the locus on chromosome 19, the association signal overlapped with several genes including *TGFB1, AXL, HNRNPUL1* and *CCDC97*, with the most significant association for the intronic *TGFB1* variant rs8105161 (EAF = 16%, OR = 0.8 [0.74-0.86], *p* = 1.76*10^−8^). In chromosome 20, the associated variants were located in the first intron of *EYA2*. In two of the significant loci, the associated variants were tightly clustered in the region of antisense genes: *COL4A2-AS2* and an intronic region of *COL4A2* in chromosome 13, and *STX-17-AS1* in chromosome 19.

The two variants most significantly associated with otosclerosis on choromosome 14, rs1951391 (EAF = 33%, OR = 1.38 [1.28-1.49], *p* = 7.0*10^−17^) and rs62007683 G>T (EAF = 33%, OR = 1.37 [1.27-1.47], *p* = 3.2*10^−16^), were both located in the same intronic region of *MARK3*. In the association locus on chromosome 6, an apparent association haplotype block spanned the entire *SUPT3H* gene and the first exon and first intron of the *RUNX2* gene; the lead variant rs13192457 (EAF = 44%, OR = 1.25 [1.17-1.34], *p* = 2.3*10^−10^) is located in an intronic region of *SUPT3H*.

### Fine-mapping of the association loci

To identify the most likely causal variants in the otosclerosis association loci, we performed a fine-mapping analysis in FinnGen using the FINEMAP software (v1.4).^25^ We analyzed ten loci that reached genome-wide significance in the meta-analysis and had suggestive evidence of association in FinnGen (*p* < 1*10^−6^). The number of variants in the resulting credible sets ranged from three (for the chromosome 4 locus near *MEPE*) to 4,604 reflecting high uncertainty (Supplementary Data 6). In the chromosome 4 locus, the *MEPE* frameshift variant rs753138805 was the most likely causal variant (probability = 66%). In the chromosome 17 locus upstream of *FAM20A*, the credible set included nine variants, with the most likely causal variants being rs11868207 (probability = 52%, AF = 23.6%, OR = 1.39 [1.29-1.49], p = 2.1*10^−18^) and rs8070086 (probability = 32%, AF = 23.6%, OR = 1.39 [1.29-1.49], p = 1.8*10^−18^).

### Gene and gene set analysis

In a gene-based analysis using MAGMA, a total of 62 genes were significantly associated with otosclerosis (*p* < 2.6*10^−6^) (Supplementary Data 7). The gene-based associations were significantly enriched (*p* < 4.9*10^−6^) for the following gene sets based on gene ontology terms: COLLAGEN TYPE IV TRIMER (*p* = 4.1*10^−7^), BASEMENT MEMBRANE COLLAGEN TRIMER (*p* = 3.7*10^−7^), TRANSFORMING GROWTH FACTOR BETA RECEPTOR ACTIVITY TYPE I (*p* = 1.7*10^−6^), POSITIVE REGULATION OF CHOLESTEROL METABOLIC PROCESS *p* = 7.9*10^−7^), and BIOMINERALIZATION (*p* = 2.2*10^−6^).

Following the identification of a genome-wide significant association signal overlapping with the *TGFB1* gene and gene-set association with type I TGFβ receptor activity, we examined other genes involved in TGFβ1 signalling for subthreshold association signals (Supplementary Data 8).^26^ We observed a trend towards association in the *TGFBR1* locus but not the *TGFBR2* locus. The lead variant in the *TGFBR1* locus, rs11386616 C>CG (EAF = 24%, OR = 1.18 [1.1-1.26], *p* = 2.0*10^−6^) is eQTL for *TGFBR1* in whole blood in the GTEx database (NES = -0.12, *p* = 2.3*10^−13^). The *TGFBR1* gene was also significantly associated with otosclerosis in the gene-based analysis using MAGMA (*p* = 1.8*10^−6^). Within the regions coding for the SMAD proteins that function as the main signal transducers of the TGFβ superfamily, we observed modest and nonsignificant association signals near *SMAD3* (intronic lead variant rs7163381 A>G [AF= 75%, OR = 0.8 [0.80-0.86], *p* = 1.0*10^−5^]) and in the region near *TGFBI* (coding for transforming growth factor-beta-induced) and *SMAD5* (lead variant: rs17169386 T>C; EAF = 0.012, OR = 2.05 [1.43-2.95], *p* = 1.3*10^−4^).

### Associations with gene expression and splicing

We examined the association of lead variants with tissue-specific gene expression in the GTEx v8 database.^23^ Among 18 lead variants, 11 variants were eQTL in at least one tissue at the 5% FDR cut-off provided by the GTEx project (Supplementary Data 11).^23^ The 11 variants were eQTL for a total of 53 genes in 43 tissues and in addition, seven lead variants were sQTL for a total of 22 genes in 42 tissues (Supplementary Data 12). Eleven genes (*TGFB1, CKB, RUNX2, AHSG, TPBG, CLCN7, SPP1, PKD2, NEAT1, LTBP3*, and *ITPR3*) have been implicated in bone metabolism or skeletal disorders, and are described in more detail in the Discussion section.

The eQTL and sQTL signals suggest a regulatory role for many of the identified otosclerosis susceptibility variants. In several loci, the lead variant was associated with the expression or splicing of multiple genes. In the chromosome 14 locus characterized by an association haplotype block spanning *MARK3* and *CKB*, the lead variant rs1951391 T>C (EAF = 33%, OR = 1.38 [1.28-1.49], *p* = 7.0*10^−17^) was associated with the expression of both *MARK3* (eQTL for 20 tissues and sQTL for 4 tissues) and *CKB* (eQTL for 7 tissues and sQTL for 8 tissues); the variant was also eQTL for nine and sQTL for five other genes in close proximity. In the chromosome 6 locus with an association block overlapping *SUPT3H* and part of *RUNX2*, the lead variant rs13192457 (EAF = 44%, OR = 1.25 [1.17-1.34], *p* = 2.3*10^−10^) was eQTL for *SUPT3H* in several tissues (e.g. NES = 0.2 in cultured fibroblasts; *p* = 9.1*10^−8^) and an sQTL for *RUNX2* in testes (NES = -0.47, *p* = 7.5*10^−15^). In the chromosome 6 locus near *IP6K3*, the lead variant rs9469583 (EAF = 58%, OR = 1.22 [1.15-1.3], *p* = 2.1*10^−10^) was most strongly associated with the expression (normalized expression score [NES] = -0.7, *p* = 8.2*10^−30^ in pancreas tissue) and splicing (NES = 0.86 in skeletal muscle, *p* = 1.1*10^−72^) of *IP6K3*; however, the variant was also eQTL and sQTL for several other genes, including *ITPR3, UQCC2* and *BAK1*.

We also sought to identify variants simultaneously associated with reduced susceptibility for otosclerosis and reduced gene expression, as such genes could represent targets for therapeutic inhibition. The intronic rs8105161 lead variant in *TGFB1* was associated with protection from otosclerosis (OR = 0.8 [0.74-0.86], *p* = 1.8*10^−8^) and reduced expression of *TGFB1* in adrenal glands (NES = -0.76, *p* = 1.3*10^−19^) and cultured fibroblasts (NES = -0.097, *p* = 1.1*10^−4^). In the chromosome 13 locus, in which the association signal clustered in the region of *COL4A2-AS2* (COL4A2 antisense 2) within the *COL4A2* gene, the lead variant rs9559805 (OR = 0.82 [0.77-0.87] for otosclerosis, *p* = 3.4*10^−11^) was associated with reduced expression of both *COL4A2-AS2* (NES = -0.17 in the muscularis layer of the esophagus, *p* = 3.5*10^−6^) and *COLA42* (NES = -0.16, *p* = 8.3*10^−5^ in subcutaneous adipose tissue).

### Replication of previous otosclerosis candidate genes

#### Candidate genes

As two of the genome-wide significant loci from the meta-analysis overlapped with candidate genes (*TGFB1* and *MEPE*), we also examined the association signals in the regions of other candidate genes (Supplementary Data 9; Supplementary Data 10). Nine candidate variants were included in the meta-analysis. Four previously reported susceptibility variants in or near the *COL1A1* gene were associated with otosclerosis at nominal significance in the meta-analysis, but only the upstream gene variant rs11327935 GA>G (EAF = 18%, OR = 1.11 [1.03-1.2], p = 7.3*10^−3^) was nominally significant after multiple correction (OR = 1.12 [1.04-1.21], *p* = 0.0041).^27-29^ None of the previously reported susceptibility variants in *SERPINF1* were included in the current meta-analysis.^13,30^ Observable candidate variants in *BMP2* and *BMP4* were not significant in the meta-analysis.^31^ However, we observed a significant association approximately 300kb upstream of *BMP2* overlapping the long non-coding RNA *CASC20*. The lead variant in this locus, rs2225347 A>T (EAF = 42%; OR = 0.82 [0.76-0.88]; *p* = 6.2*10^−8^), is not reported as eQTL or sQTL for *BMP2* or any gene in the GTEx v8 database.^23^

#### Linkage loci

Among published linkage loci, one (OTSC7, 6q13–16.1) has been narrowed to a region containing only 12 candidate genes.^32^ In this region, we observed a trend towards association for variants near the gene *CD109*, with the lowest p-value for the intronic *CD109* variant rs10805974 G>C (AF= 36%, OR = 1.16 [1.09-1.23], *p* = 3.4*10^−6^). *CD109* was significantly associated with otosclerosis in the gene-based analysis using MAGMA (*p* = 1.2*10^−8^).

### Further associations with the 18 otosclerosis lead variants

#### Phenome-wide association studies

To assess pleiotropic effects of genetic risk factors to otosclerosis, we first examined the phenome-wide associations of the 18 lead variants from the meta-analysis with a total of 2419 traits in UKBB and 2925 traits in FinnGen. Six lead variants were associated with standing height, and five with heel bone mineral density (BMD) in UKBB (*p* < 2.1*10^−5^) (Supplementary Data 13). The intronic *MARK3* lead variant rs1951391 was associated with forearm fractures in FinnGen (*p* = 3.8*10^−6^) and the rs753138805 frameshift variant in *MEPE* was associated with lower leg fractures (*p* = 3.8*10^−6^).

#### GWAS Catalog

Among the 18 lead variants, three were reported in the GWAS Catalog as being associated with either height, BMD or BMI-adjusted waist-hip ratio (Supplementary Data 13).^33^ Overall, among all the 1,257 genome-wide significant variants associated with otosclerosis in our meta-analysis, 62 variants have been reported to be associated with a total of 45 traits (Supplementary Data 14). Eight loci have been associated with BMD, four with height, and two with osteoarthritis. Variants associated with increased risk of otosclerosis were most often associated with increased height and decreased BMD. Additionally, in the chromosome 8 locus near the *EIF3H* gene, an intergenic variant associated with otosclerosis at genome-wide significance (rs13279799, OR = 0.77 [0.71-0.83]; *p* = 2.8*10^−11^) has been previously associated with ossification of the longitudinal ligament of spine, a rare form of heterotopic ossification.^34^

#### Shared heritability

To interrogate shared heritability between the traits, we calculated pairwise genetic correlations (r_g_) using summary statistics from the otosclerosis meta-analysis and published large GWA studies.^20,35,36^ Otosclerosis was genetically correlated with fracture risk (r_g_ = 0.24 [S.E. 0.08], *p* = 0.0034) and height (r_g_ = 0.10 [S.E. 0.04], *p* = 0.0177), but not with BMD (r_g_ = -0.088 [S.E. 0.056], *p* = 0.12).

## Discussion

Although otosclerosis is highly heritable its genetic background is still poorly understood. Previous studies have reported one GWAS locus (*RELN*), while results for candidate genes have been inconclusive. Here, we identify 18 loci associated with otosclerosis, of which 15 are novel. We also replicate the *RELN* locus and two candidate gene loci – *TGFB1* and *MEPE*. These results offer further insights to disease pathophysiology.

We confirm a polygenic basis to otosclerosis and identify loci harbouring potentially causative genes involved in the regulation of bone structure. Earlier studies assumed a dominant inheritance, but the inability to map otosclerosis to one or a few high-impact genes has suggested a complex nature. Many etiologies to otosclerosis have been proposed, including differences in TGFβ, parathyroid hormone or angiotensin II signalling, alterations in collagen type I, inflammation, viral disease, and autoimmunity.^37^

A considerable number of association loci are associated with severe skeletal disorders (Table 3). Disordered or heterotopic bone growth is a common feature e.g. in diaphyseal dysplasia (caused by mutations in *TGFB1*), osteopetrosis subtypes (caused by mutations in *CLCN7*), and ossification of the posterior longitudinal ligament of spine (GWAS signal for rs13279799 in the chromosome 8 locus). Otosclerosis presents later in adulthood and represents a more limited and common phenotype compared with severe skeletal dysplasias. Shared heritability with height and appendicular fracture risk across the genome also suggests more cumulative effects on the regulation of bone growth and remodeling.

**Table 3:**
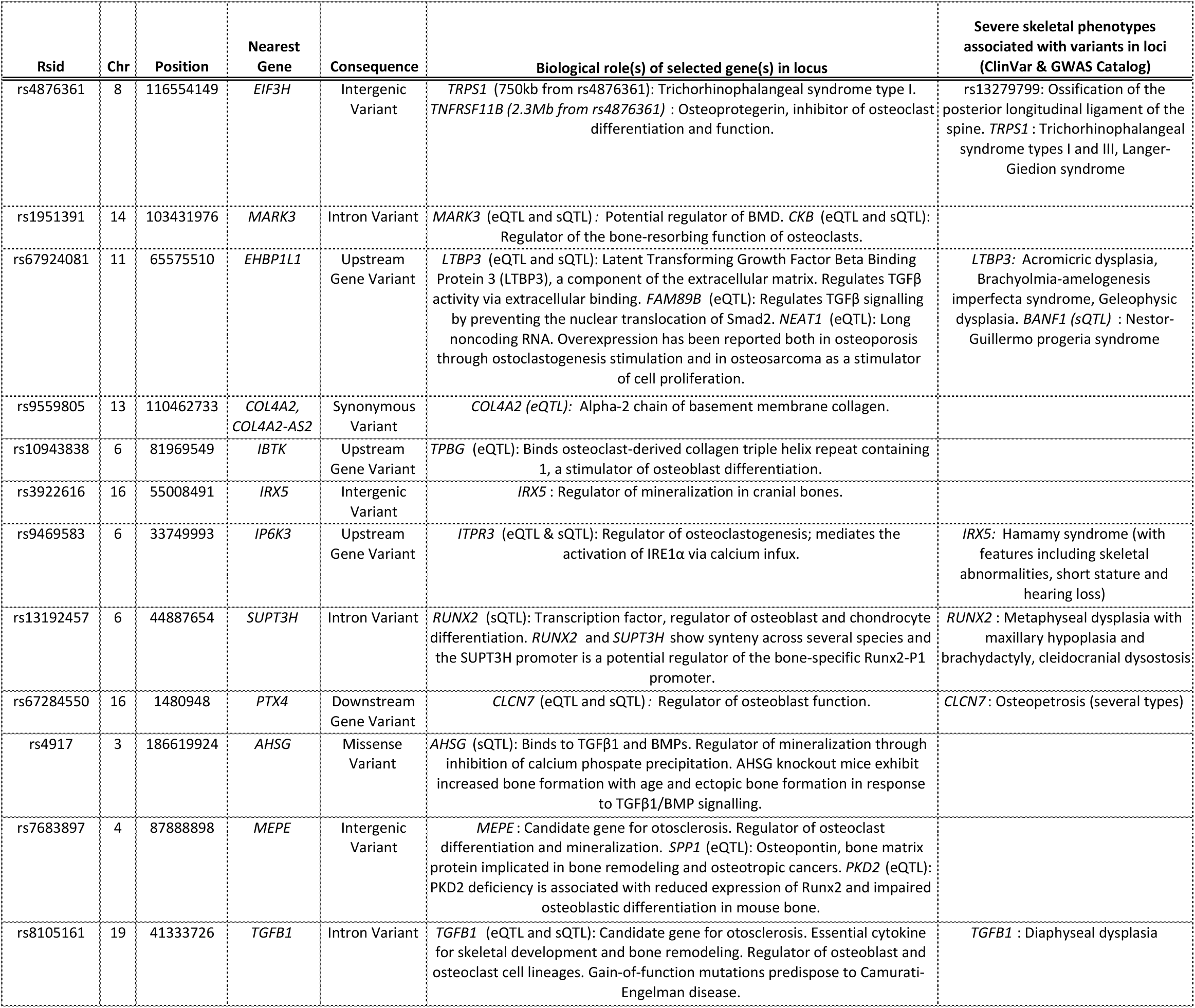
Biological functions of genes in selected association loci. In 13 association loci, genes implicated in bone metabolism are in close proximity with the lead variant or associated with the expression of the lead variant. eQTL = expression quantitative trait locus, sQTL = splicing quantitative trait locus. References to individual studies are listed in Supplementary Data 15.

The effects on bone growth and remodeling are likely mediated by several pathways. The known biological functions of many putative risk genes in otosclerosis association loci include regulation of osteoblasts – the cells responsible for bone formation – and osteoclasts – responsible for the breakdown of bone, either through effects on cell differentiation or function (Table 3). Some genes (*AHSG, IRX5*, and *MEPE*) are also directly involved in biomineralization, and *AHSG* and the candidate gene *MEPE* have been reported to have dual roles in the regulation of mineralization and osteoclast or osteoblast cell lineages.

Recently, Shrauwen and colleagues proposed a model of otosclerosis in which increased bone turnover results from mutations ablating two functional motifs of *MEPE*, leading to both accelerated osteoclast differentiation and enhanced mineralization and thus increased bone turnover.^38^ In contrast with otosclerosis, they observed frameshift mutations ablating only one functional motif of *MEPE* in individuals with a severe craniofacial defect with thickening of the skull, potentially reflecting an unopposed increase in mineralization. Here, we replicate at genome-wide significance an association between otosclerosis and a frameshift mutation that is likely to ablate both functional motifs in *MEPE*. Reflecting a systemic effect on the skeleton, the rs753138805 variant is also associated with increased leg fracture risk, similarly to an unpublished study in the Norwegian HUNT cohort.^39^

Several genes in the susceptibility loci converge on Transforming growth factor beta-1 (TGFβ1) signalling pathways. *TGFB1* has long been proposed as a susceptibility gene for otosclerosis, but the quality of the supporting evidence has been relatively weak.^15-18,28,40^ Here, for the first time, we report an association at the *TGFB1* locus at genome-wide significance, with the strongest association for the intronic variant rs8105161. The significance of TGFβ1 signalling is supported by gene set enrichment and analysis of other loci.

TGFβ1, a member of the transforming growth factor beta superfamily, is a cytokine with an essential role in skeletal development and mature bone remodeling, regulating both osteoblast and osteoclast cell lineages.^41,42^ Gain-of-function mutations in *TGFB1* predispose to diaphyseal dysplasia, and mutations in other TGFβ superfamily members can cause multiple skeletal disorders of varying severity.^43^

Other potentially causative TGFβ-related genes in the otosclerosis susceptibility loci include *RUNX2* in chromosome 3, *AHSG* in chromosome 8, and *LTBP3* and *FAM89B* in chromosome 11, all of which are associated with the expression of the respective lead variants. RUNX2 is a transcription factor regulated by TGFβ1-signalling via both the canonical Smad and p38 MAPK pathways and has an essential role in osteoblast and chondrocyte differentiation.^41,42,44^Alpha2-HS-glycoprotein/fetuin (AHSG) antagonizes TGFβ1-signalling by binding to TGFβ1 and TGFβ related bone morphogenic proteins (BMPs).^45,46^ It can also affect mineralization directly by inhibiting calcium phosphate precipitation via formation of calcium-fetuin complexes.^47-49^ LTBP3 can regulate the latency and activation of TGFβ1 through direct extracellular binding.^50^ The nearby *FAM89B* codes for Leucine repeat adapter protein 25, which may regulate TGFβ signaling by preventing the nuclear translocation of Smad2.^51^

In addition to signaling proteins, *COL1A1* coding for the major subunit of type I collagen is a prominent candidate gene for otosclerosis.^12,27-29,52^ Mutations in *COL1A1* predispose to osteogenesis imperfecta, often also characterized by hearing loss. In osteogenesis imperfecta, similarly to carriers of truncating *MEPE* variants, increased bone turnover in the middle could occur in association with general skeletal fragility. While we do not observe a strong signal for *COL1A1*, similarly to most examined candidate genes, we report an intronic association signal in *COL4A2*, coding for a subunit of collagen IV, located in the basement membrane. Future mechanistic studies could examine whether *COL4A2* has a structural or signaling role in otosclerosis. *COL4A2* is highly conserved across species, and its mutations have been reported in a broad spectrum of organ anomalies, but we are not aware of a previous association with skeletal disorders.^53^ Although type IV collagens regulate BMPs in Drosophila, a similar role in humans is uncertain.^54^

Our results highlight several genes and signaling pathways for follow-up mechanistic studies. Genetic discovery has also increasingly preceded or guided therapeutic development. Future sequencing studies could aim to discover loss-of-function mutations within the susceptibility loci to approximate the effects of therapeutic inhibition. In the case of *TGFB1*, loss-of-function mutations are exceedingly rare and functional studies are needed to assess the effect of direct TGFβ1 inhibition.^24^ Of note, an inhibitor of BMP type I receptor kinases has been studied for the treatment of ectopic ossification; however, in our meta-analysis, its targets (coded by *ACVR1* and *BMPR1A)* were not significantly associated with otosclerosis.^55^ Reflecting the heterogeneity of conditions associated with different TGFβ superfamily genes, therapeutic intervention may need to be precisely tailored to each condition.^41^

Although we present the largest GWAS study of otosclerosis to date, our study has limitations. The identification of otosclerosis cases is based on ICD diagnoses, which we could not verify with audiometric or imaging data. However, the prevalence of stapes procedures among cases is concordant with clinical experience. As bone data are not available within the GTEx consortium, the analyses of gene expression rely on other tissues; associations in bone could differ. The physical proximity of lead variants with biologically relevant genes does not prove causation; identified and unidentified variants in the loci can exert their effects through yet unknown mechanisms.

In summary, we determine a polygenic basis to otosclerosis with 18 genome-wide significant susceptibility loci, and shared heritability between otosclerosis and other common and rare skeletal traits. The loci offer multiple potential avenues for understanding disease pathophysiology through genes involved in bone remodeling, mineralization, and basement membrane collagen composition. In particular, our results highlight several TGFβ superfamily members for follow-up studies.

## Methods

### Cohort descriptions

We identified individuals with ICD-based otosclerosis diagnoses from three national biobank-based cohorts: The Finnish FinnGen cohort, the Estonian Biobank (EstBB) and the UK Biobank (UKBB).

The FinnGen data used here comprise 210,932 individuals from FinnGen Data Freeze 5 (https://www.finngen.fi). The data were linked by unique national personal identification numbers to the national hospital discharge registry (available from 1968) and the specialist outpatient registry (available from 1998). Data comprised in FinnGen Data Freeze 5 are administered by regional biobanks (Auria Biobank, Biobank of Central Finland, Biobank of Eastern Finland, Borealis Biobank, Helsinki Biobank, Tampere Biobank), the Blood Service Biobank, the Terveystalo Biobank, and biobanks administered by the Finnish Institute for Health and Welfare (THL) for the following studies: Botnia, Corogene, FinHealth 2017, FinIPF, FINRISK 1992–2012, GeneRisk, Health 2000, Health 2011, Kuusamo, Migraine, Super, T1D, and Twins). The FinnGen study protocol (HUS/990/2017) was approved by the Ethics Review Board of the Hospital District of Helsinki and Uusimaa.

EstBB is a population-based cohort of 200,000 participants with a rich variety of phenotypic and health-related information collected for each individual.^56^ At recruitment, participants have signed a consent to allow follow-up linkage of their electronic health records (EHR), thereby providing a longitudinal collection of phenotypic information. EstBB allows access to the records of the national Health Insurance Fund Treatment Bills (from 2004), Tartu University Hospital (from 2008), and North Estonia Medical Center (from 2005). For every participant there is information on diagnoses in ICD-10 coding and drug dispensing data, including drug ATC codes, prescription status and purchase date (if available).

UKBB comprises phenotype data from 500,000 volunteer participants from the UK population aged between 40 and 69 years during recruitment in 2006-2010.^57^ Data for all participants have been linked with national Hospital Episode Statistics.

### Identification of otosclerosis cases

Case status was assigned for individuals with any ICD-10 H80* code, or when available, the ICD-9 code 387 or ICD-8 code 386 (Table 1). In FinnGen, only ICD codes registered in specialty care settings were used for case definition. Stapes procedures were identified based on the Nomesco codes DDA00 (stapedotomy) and DDB00 (stapedectomy) in FinnGen and the national health insurance treatment service code 61006 (stapedotomy) in EstBB.

### Genotyping and imputation of variants

FinnGen samples were genotyped using Illumina and Affymetrix arrays (Illumina Inc., San Diego, and Thermo Fisher Scientific, Santa Clara, CA, USA). Genotype imputation was performed using a population-specific SISu v3 imputation reference panel comprised of 3,775 whole genomes as described (https://www.protocols.io/view/genotype-imputation-workflow-v3-0-xbgfijw).

The samples from the Estonian Biobank were genotyped at the Genotyping Core Facility of the Institute of Genomics, University of Tartu using the Global Screening Array (GSAv1.0, GSAv2.0, and GSAv2.0_EST) from Illumina. Altogether 155,772 samples were genotyped and PLINK format files were exported using GenomeStudio v2.0.4.^58^ Individuals were excluded from the analysis if their call-rate was < 95% or if the sex defined based on heterozygosity of the X chromosome did not match the sex in phenotype data. Variants were excluded if the call-rate was < 95% and HWE p-value < 1e-4 (autosomal variants only). Variant positions were updated to genome build 37 and all alleles were switched to the TOP strand using tools and reference files provided at https://www.well.ox.ac.uk/∼wrayner/strand/. After QC the dataset contained 154,201 samples for imputation. Before imputation variants with MAF<1% and indels were removed. Prephasing was done using the Eagle v2.3 software.^59^ The number of conditioning haplotypes Eagle2 uses when phasing each sample was set to: --Kpbwt=20000. Imputation was done using Beagle v.28Sep18.793 with effective population size ne=20,000.^60^ An Estonian population specific imputation reference of 2297 WGS samples was used.^61^

UKBB samples have been genotyped and imputed as described previously.^57^ Additional quality controls of version 3 data have been performed and publicly reported by the Pan-UKB team from the Analytic & Translational Genetics Unit (ATGU) at Massachusetts General Hospital and the Broad Institute of MIT and Harvard (https://pan.ukbb.broadinstitute.org.2020). The analysis referenced in this study is based on samples from 419,622 individuals with European ancestry.

### Statistical analysis

GWAS for the individual study cohorts was performed using a generalized mixed model with the saddlepoint approximation using SAIGE v0.20, using a kinship matrix as a random effect and covariates as fixed effects.^62^ For all cohorts, variants with a minor allele frequency less than 0.1% were excluded.

In FinnGen, samples from individuals with non-Finnish ancestry and twin/duplicate samples were excluded, and GWAS was performed for 1,350 cases and 209,582 controls. Age, sex, 10 PCs and the genotyping batch (for batches with at least 10 cases and controls) were used as covariates. Results were filtered to variants with imputation INFO score > 0.6. In EstBB, a GWAS was performed on 713 cases and 136,7933 controls of European ancestry including related individuals and adjusting for the first 10 PCs of the genotype matrix, as well as for birth year and sex. The statistical analysis of UKBB data has been performed and publicly reported by the Pan-UKB team (https://pan.ukbb.broadinstitute.org.2020). The UKBB GWAS results are reproduced here for selected variants for the purpose of cross-cohort replication and inclusion in the meta-analysis.

Variant positions in the UKBB and EstBB summary statistics were lifted from GRCh37 to GRCh38 using the liftOver v1.12 Bioconductor R package.^63^ To assess similarity of genetic effects between cohorts, we used the cross-trait LD Score Regression to calculate pairwise genetic correlations (r_g_) based on summary statistics from each cohort.^20,21^ Summary statistics from the individual cohorts for 13,762,819 variants present in at least two cohorts with a cross-cohort minor allele frequency > 0.1% and imputation INFO score > 0.7 were combined using an inverse-variance weighted fixed-effect meta-analysis with GWAMA v2.2.2.^64^ The following analyses are based on summary statistics from the meta-analysis unless otherwise specified.

### Characterization of association loci

For the individual cohorts and meta-analysis, we merged genome-wide significant variants within 500kb of each other into association loci. The lead variants from these loci were not in LD with each other based on individual-level data from FinnGen (r^2^ < 1*10^−4^ for all variant pair comparisons). We annotated the lead variants by mapping their physical position to genes and consequences using the Ensembl Variant Effect Predictor (VEP) based on the GRCh38 genome build.^65^ In addition, we annotated all genome-wide significant variants and all variants in high LD (r>0.6) with the lead variants from the meta-analysis using VEP. As the highest number of association loci was observed in FinnGen, we estimated LD in the FinnGen cohort using PLINK v1.07.^66,67^ LocusZoom was used to visualize the association loci and the regions surrounding other genes of interest.^68^

### Fine-mapping

Based on otosclerosis summary statistics from FinnGen, we fine-mapped all regions where the lead variant reached a p-value of < 1*10^−6^ in FinnGen using FINEMAP v1.4.^25^ We used a 3Mb window (± 1.5Mb) around each lead variant, and calculated LD between each variant from individual-level FinnGen data. For credible sets with over 10 variants, we report those with a ≥ 0.01 probability of being causal.

### eQTL and sQTL mapping

We used the GTEx v8 database (https://gtexportal.org) to map otosclerosis susceptibility variants to genes and tissues.^23^ We downloaded the data for all 49 tissues, and mapped the lead variants from the meta-analysis, as well as selected variants of interest (e.g. from candidate gene loci), to eQTL (expression quantitative locus) or sQTL (splicing quantitative locus) associations using the 5% false discovery rate cut-off provided by the GTEx Consortium.

### Gene and gene set -based analyses

We used MAGMA v1.07 to identify genes and gene sets associated with otosclerosis based on effect estimates from the meta-analysis.^69^ Variants were mapped to 18,910 genes based on their RefSNP numbers. We performed a gene-based analysis using the default SNPwise-mean model and a Bonferroni-corrected *p*-value threshold (α = 0.05/18,910). Based on the results from the gene-based analysis, we then performed a competitive gene set based analysis using 10,182 GO term based gene sets downloaded from the Molecular Signature Database v7.1, using a Bonferroni-corrected *p*-value threshold (α = 0.05/10,182).^70^

### Phenome-wide association analyses and genetic correlations

We examined the pleiotropic effects of lead variants based on phenome-wide association studies in FinnGen and UKBB. In FinnGen, GWAS was performed for 2,925 registry-based disease endpoints similarly to the GWAS for otosclerosis. The study participants were linked with national registries covering the whole population for hospital discharges (data available since 1968), deaths (1969–), outpatient specialist appointments (1998–), cancers (1953–), and medication reimbursements (1995–). Disease endpoints were collated based on International Classification of Diseases (ICD) codes (revisions 8–10), International Classification of Diseases for Oncology (ICD-O) third edition codes, NOMESCO procedure codes, Finnish-specific Social Insurance Institute (KELA) drug reimbursement codes, and drug-specific ATC-codes. In addition, specific clinical endpoints combining relevant comorbidities and exclusion criteria have been curated in coordination with clinical expert groups. Currently available FinnGen phenotype definitions are available online at https://www.finngen.fi/en/researchers/clinical-endpoints.

For UKBB, we downloaded publicly available association results for 2419 phenotypes including diseases and anthropometric traits (http://www.nealelab.is/uk-biobank & http://pheweb.sph.umich.edu:5000). Bonferroni-corrected *p*-value thresholds were used for each cohort (α = 0.05/2925 for FinnGen and α = 0.05/2419 for UKBB). Additionally, using the GWAS Catalog database (accessed on 22 April 2020), we performed a lookup of previously published association results for all variants significantly associated with otosclerosis in the meta-analysis.^33^

For three frequently occurring coassociations (height, bone mineral density (BMD), and fracture risk, we estimated the genetic correlations of each trait with otosclerosis using LD Score Regression.^20,21^ We used summary statistics from recently published large GWA studies: the GWAS meta-analysis of height in approximately 700,000 individuals by the GIANT Consortium, and the GWAS of ultrasound-quantified heel BMD and fracture risk in approximately 460,000 UKBB participants by the GEFOS Consortium.^20,35,36^

## Supporting information

Description of Supplementary Data Files

## Data Availability

FinnGen results are subjected to one year embargo and, after that, can be made available to the larger scientific community. Data are released twice a year. For more information, see: https://www.finngen.fi/en/access_results.
Genotype and phenotype data are available from the Estonian Biobank (https:// genomics.ut.ee/en/biobank.ee/data-access) upon request. 
Data from the UK Biobank (UKBB) are made available for approved researchers undertaking health research in the public good. Genomic analyses in the UKBB have been conducted and publicly reported by the Pan UKBB team under proposal number 31063. More information can be found at: https://pan.ukbb.broadinstitute.org and https://registry.opendata.aws/broad-pan-ukb/.

https://www.ukbiobank.ac.uk/scientists-3/genetic-data/

## Acknowledgements

This research has been conducted using the UK Biobank Resource under Application Number 22627. The Genotype-Tissue Expression (GTEx) Project was supported by the Common Fund of the Office of the Director of the National Institutes of Health, and by NCI, NHGRI, NHLBI, NIDA, NIMH, and NINDS. The data used for the gene expression analyses described in this manuscript were obtained from the GTEx Portal in July 2020. We thank the Pan UKBB analysis team from the Analytic & Translational Genetics Unit (ATGU) at Massachusetts General Hospital and the Broad Institute of MIT and Harvard for making UKBB summary statistics publicly available. We thank all study participants for their generous contributions to the biobanks.

