## Supplementary material for "Genome-wide Screen of Otosclerosis in Population Biobanks: 18 Loci and Shared Heritability with Skeletal Structure": Description of Supplementary Data Files

**File Name:** Supplementary Data 1. Cohort-specific Manhattan plots.

**Description:** Manhattan plots are presented for the case-control GWAS of otosclerosis in a) FinnGen (1,350 cases and 209,582 controls), b) EstBB (713 cases and 136,793 controls), and c) UKBB (350 cases and 416,007 controls). In the Manhattan plots, chromosomal positions are indicated on the x-axis and -log_10_(p-value) is presented on the y-axis for each variant. Variants with a minor allele frequency > 0.1% and imputation INFO score > 0.7 were included in the analysis.

**File Name:** Supplementary Data 2. Cohort -specific lead variants.

**Description:** Lead variants are presented for the GWAS in each cohort, annotated with effect estimates, effect allele frequencies and *p*-values in the other two cohorts. *Chr = chromosome,* *EA = effect allele, EAF = effect allele frequency, NEA = non-effect allele, OR = odds ratio, CI = confidence interval.*

**File Name:** Supplementary Data 3. Lead variants from the meta-analysis with cohort-level estimates.

**Description:** Lead variants from the meta-analysis (including a total of 2,413 cases and 762,382 controls) are annotated with effect estimates, effect allele frequencies and *p*-values within each cohort. *Chr = chromosome,* *EA = effect allele, EAF = effect allele frequency, NEA = non-effect allele, OR = odds ratio, CI = confidence interval.*

**File Name:** Supplementary Data 4. LocusZoom plots for significant loci from the meta-analysis.

**Description:** LocusZoom plots for each significant locus are shown in the same order as in Table 2. Linkage disequilibrium is shown in colour scale with the lead variants as reference, based on publicly available European genome data from the 1000 Genomes Project.

**File Name:** Supplementary Data 5. Significantly associated missense variants):

**Description:** Missense variants which were significantly associated with otosclerosis (p < 5*10^-8^) in the meta-analysis are shown. The Variant Effect Predictor tool was used for variant annotation.

**File Name:** Supplementary Data 6. FINEMAP credible sets in FinnGen.

**Description:** All regions in which the lead reached a p-value of < 1*10-6 in FinnGen were fine-mapped using FINEMAP v1.4.68. A 3Mb window (± 1.5Mb) was used around each lead variant, and LD was calculated between each variant from individual-level FinnGen data. For credible sets with over 10 variants, only variants with a ≥ 0.01 probability of being causal are reported.

**File Name:** Supplementary Data 7. Significant genes from a gene set analysis with MAGMA.

**Description:** The MAGMA v1.07 software was used to identify genes associated with otosclerosis based on effect estimates from the meta-analysis. Variants were mapped to 18,910 genes based on their RefSNP numbers. A gene-based analysis was performed using the default SNPwise-mean model and a Bonferroni-corrected p-value threshold (α = 0.05/18,910).

**File Name:** Supplementary Data 8. LocusZoom plots for genes involved in TGFβ signalling

**Description:** Data are shown for selected genes involved in TGFβ signalling, including *TGFB1*, *TGFB2*, *TGFB3*, the receptors *TGFBR1* and *TGFBR2*, and genes coding for downstream signaling proteins.

**File Name:** Supplementary Data 9. LocusZoom plots for candidate gene regions.

**Description:** LocusZoom plots are presented for the regions of four candidate genes from previous studies (*COL1A1*, *BMP2*, *BMP4*, and *SERPINF1*) and for the region of the OTSC7 linkage locus.

**File Name:** Supplementary Data 10. Replication of candidate variants.

**Description:** Meta-analysis Effect estimates and *p*-values are shown for variants previously suggested to be associated with otosclerosis. References to the original studies are presented in the main text.

**File Name:** Supplementary Data 11. eQTL for lead variants from the meta-analysis.

**Description:** We used the GTEx v8 database (https://gtexportal.org) to map otosclerosis susceptibility variants to genes and tissues. We downloaded the data for all 49 tissues, and mapped the lead variants from the meta-analysis eQTL (expression quantitative locus) associations using the 5% false discovery rate cut-off provided by the GTEx Consortium.

**File Name:** Supplementary Data 12. sQTL for lead variants from the meta-analysis.

**Description:** We used the GTEx v8 database (https://gtexportal.org) to map otosclerosis susceptibility variants to genes and tissues. We downloaded the data for all 49 tissues, and mapped the lead variants from the meta-analysis sQTL (splicing quantitative locus) associations using the 5% false discovery rate cut-off provided by the GTEx Consortium.

**File Name:** Supplementary Data 13. Phenome-wide association study of lead variants from the meta-analysis.

**Description:** Phenome-wide association study results and previous GWAS reports are presented for the lead variants from the meta-analysis. In FinnGen, GWAS was performed for 2925 registry-based disease endpoints similarly to the GWAS for otosclerosis. For UKBB, publicly available association results for 2419 phenotypes including diseases and anthropometric traits (<http://www.nealelab.is/uk-biobank> & <http://pheweb.sph.umich.edu:5000>). Bonferroni-corrected *p*-value thresholds were used for each cohort (α = 0.05/2925 for FinnGen and α = 0.05/2419 for UKBB). A search was conducted in the GWAS Catalog database (accessed on 22 April 2020) for previously published association results. *PheWAS = phenome-wide association study, GWAS = genome-wide association study.*

**File Name:** Supplementary Data 14. Previous GWAS reports for all significant variants from the meta-analysis

**Description:** Using the GWAS Catalog database (accessed on 22 April 2020), previous GWAS results were accessed for all 1,257 variants which were significantly associated with otosclerosis (p < 5*10^-8^) in the meta-analysis. *GWAS = genome-wide association study.*

**File Name:** Supplementary Data 15. References for the reported biological functions of selected genes in the association loci.

**Description:** References to the biological functions summarized in Table 3.
