## Supplementary figures and images for "Genome-wide Screen of Otosclerosis in Population Biobanks: 18 Loci and Shared Heritability with Skeletal Structure"

### Supplementary Data 1

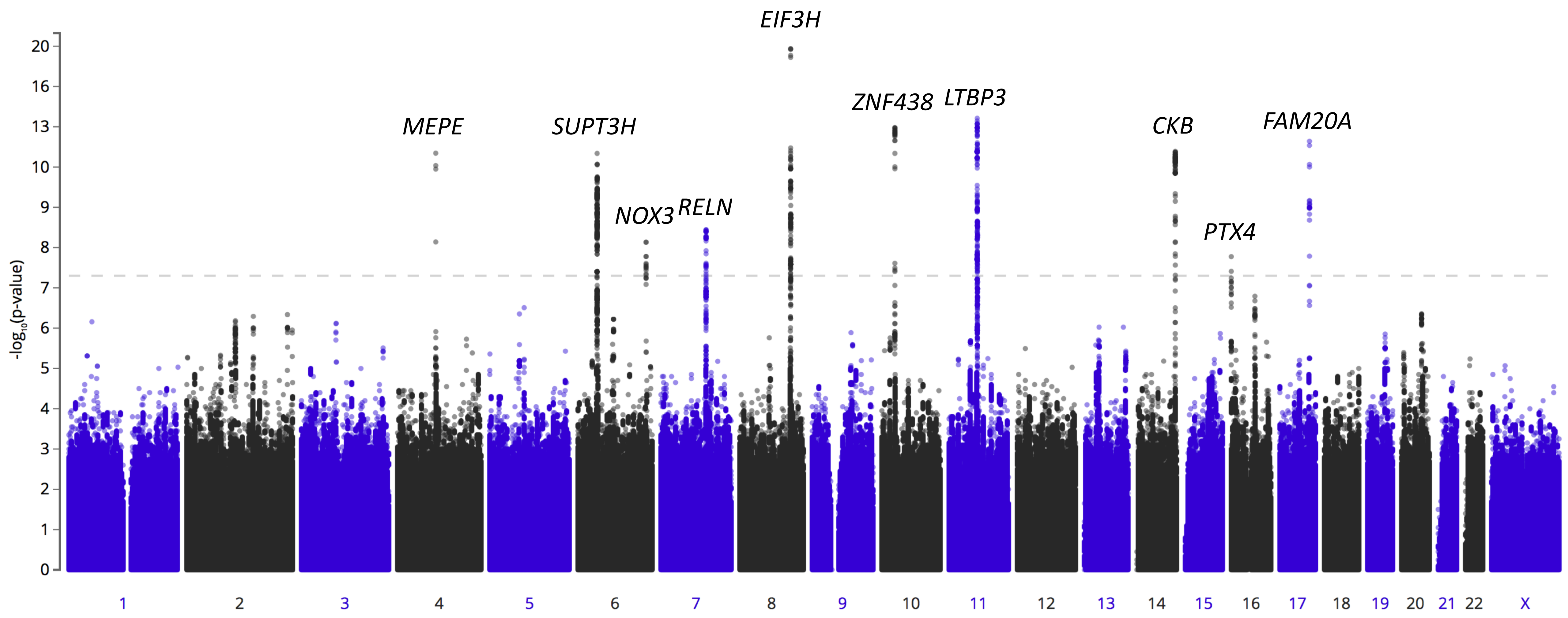

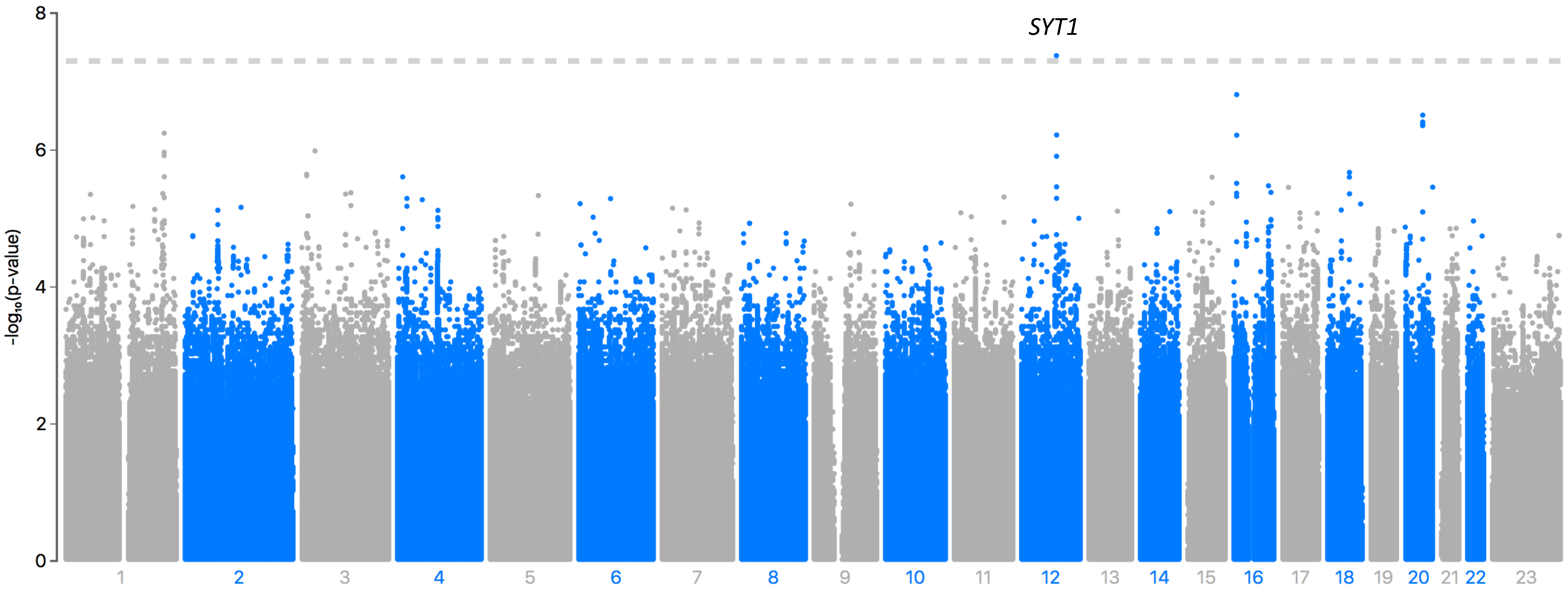

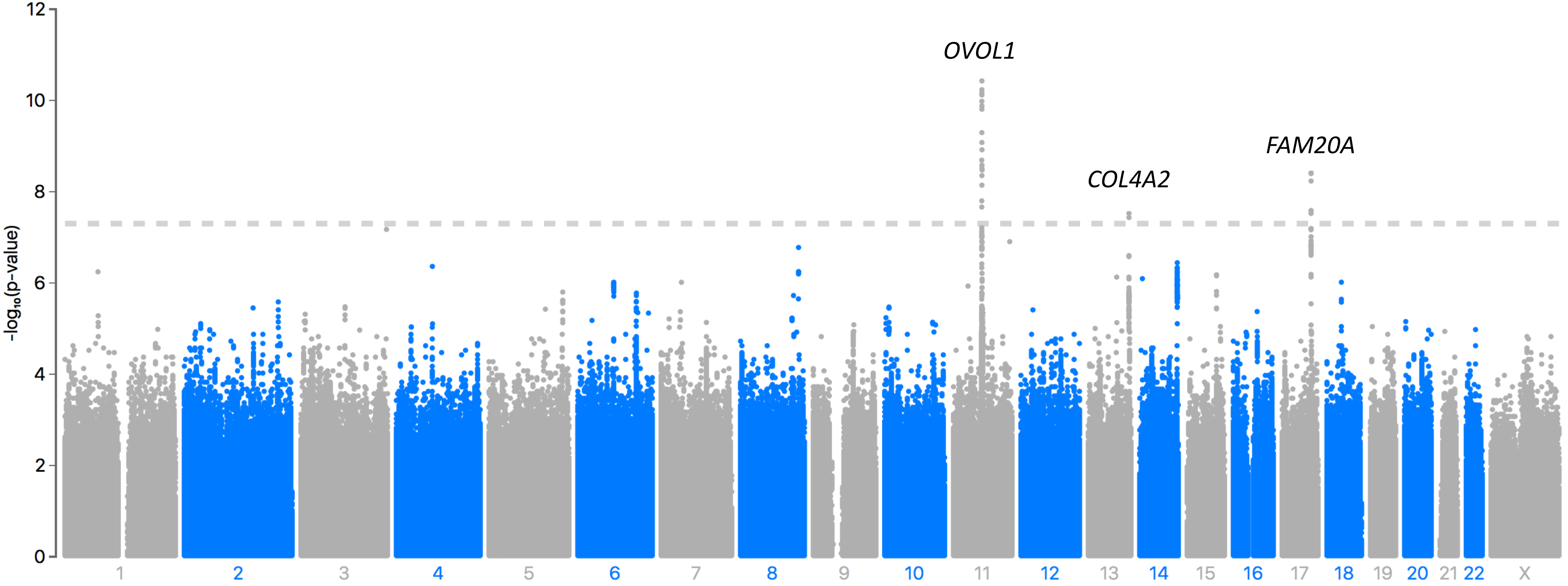

### Supplementary Data 4

#### *EIF3H* locus

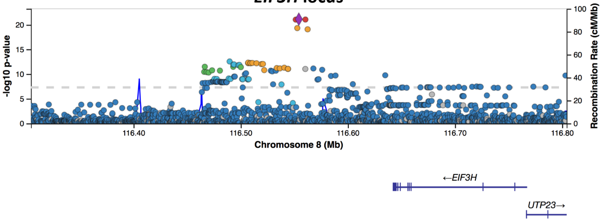


#### *LINC01482 / FAM20A* locus

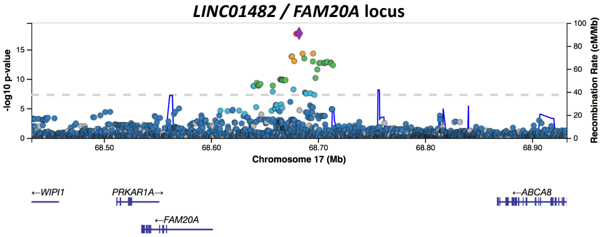


#### *MARK3* locus

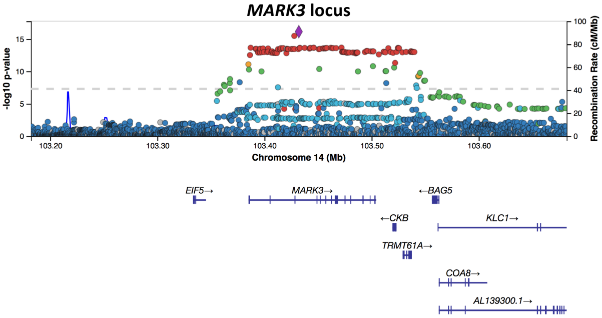


#### *ZNF438* locus

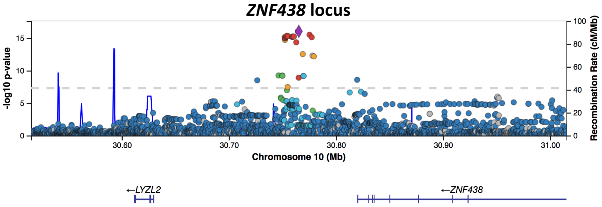


#### *EHBP1L1* locus

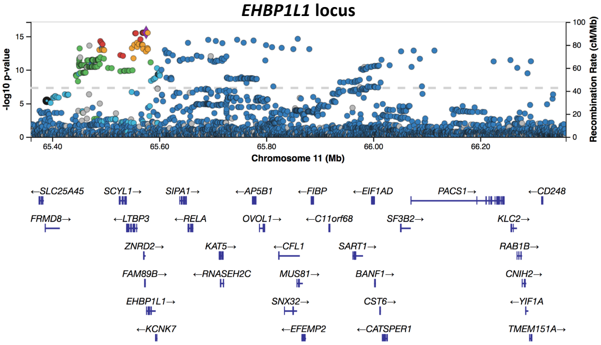


#### *RELN* locus

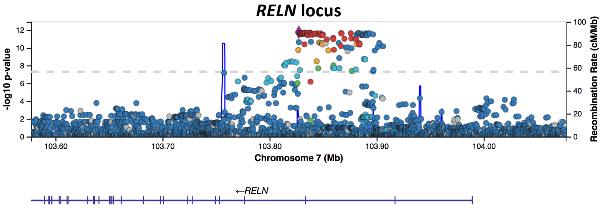

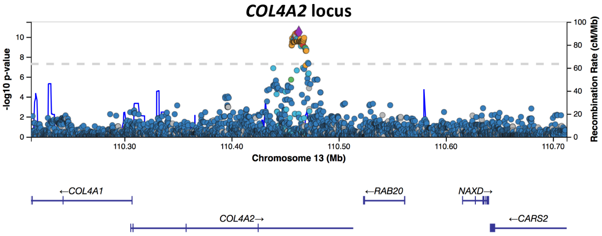

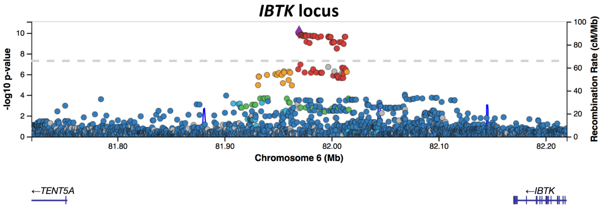

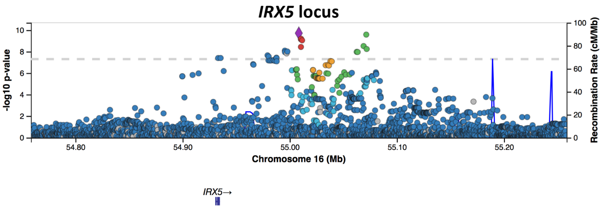

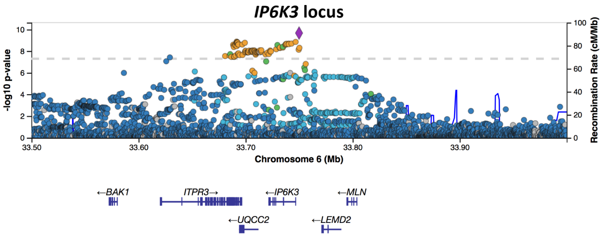

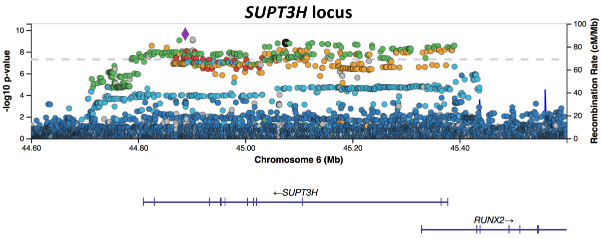

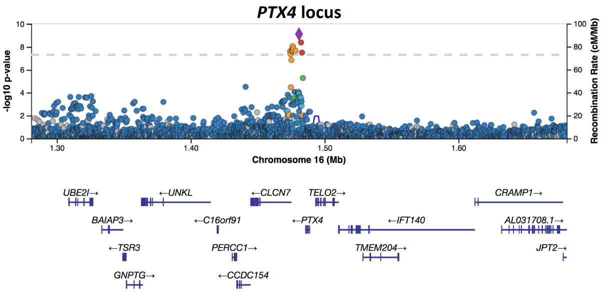

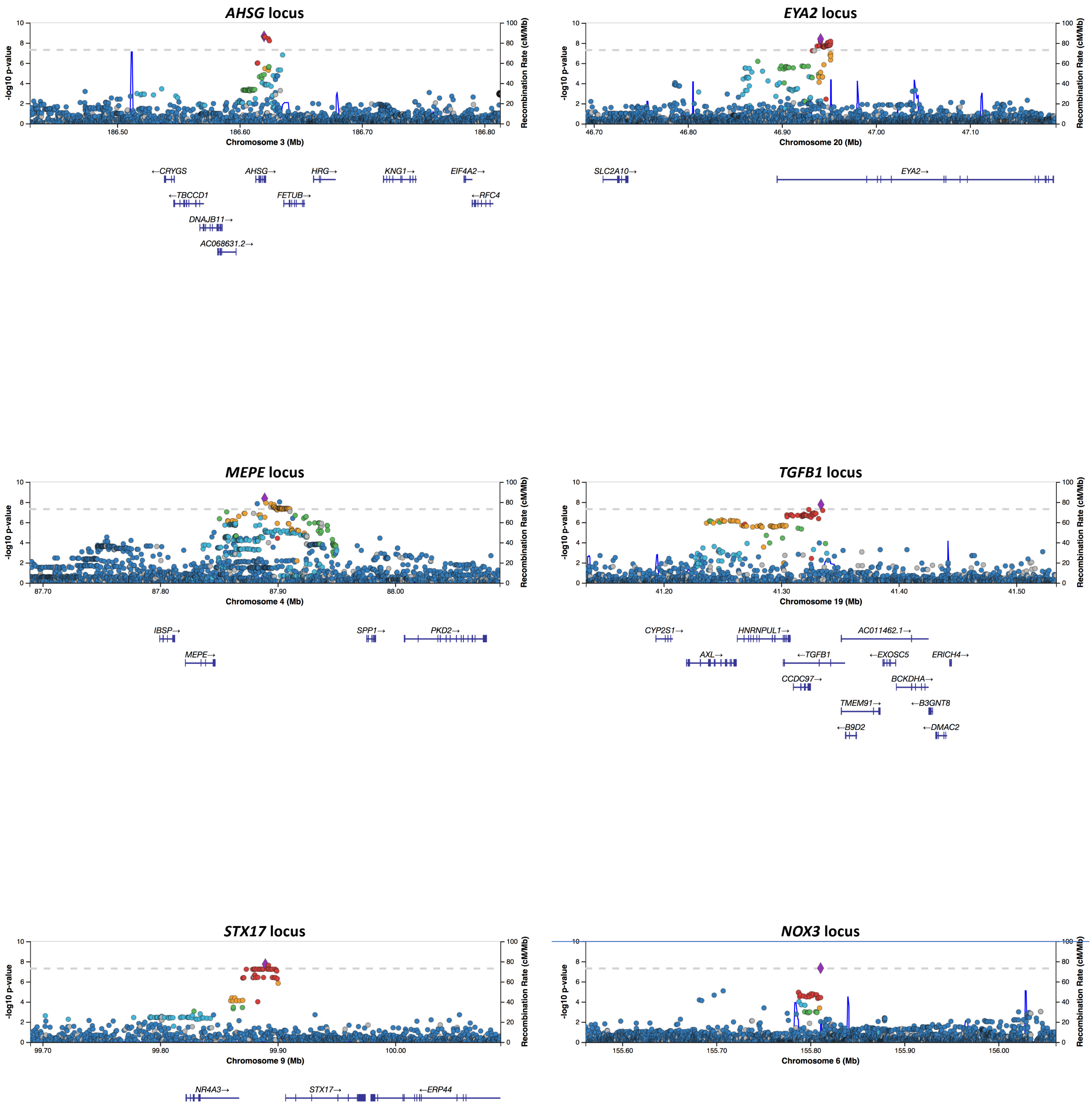

### Supplementary Data 8

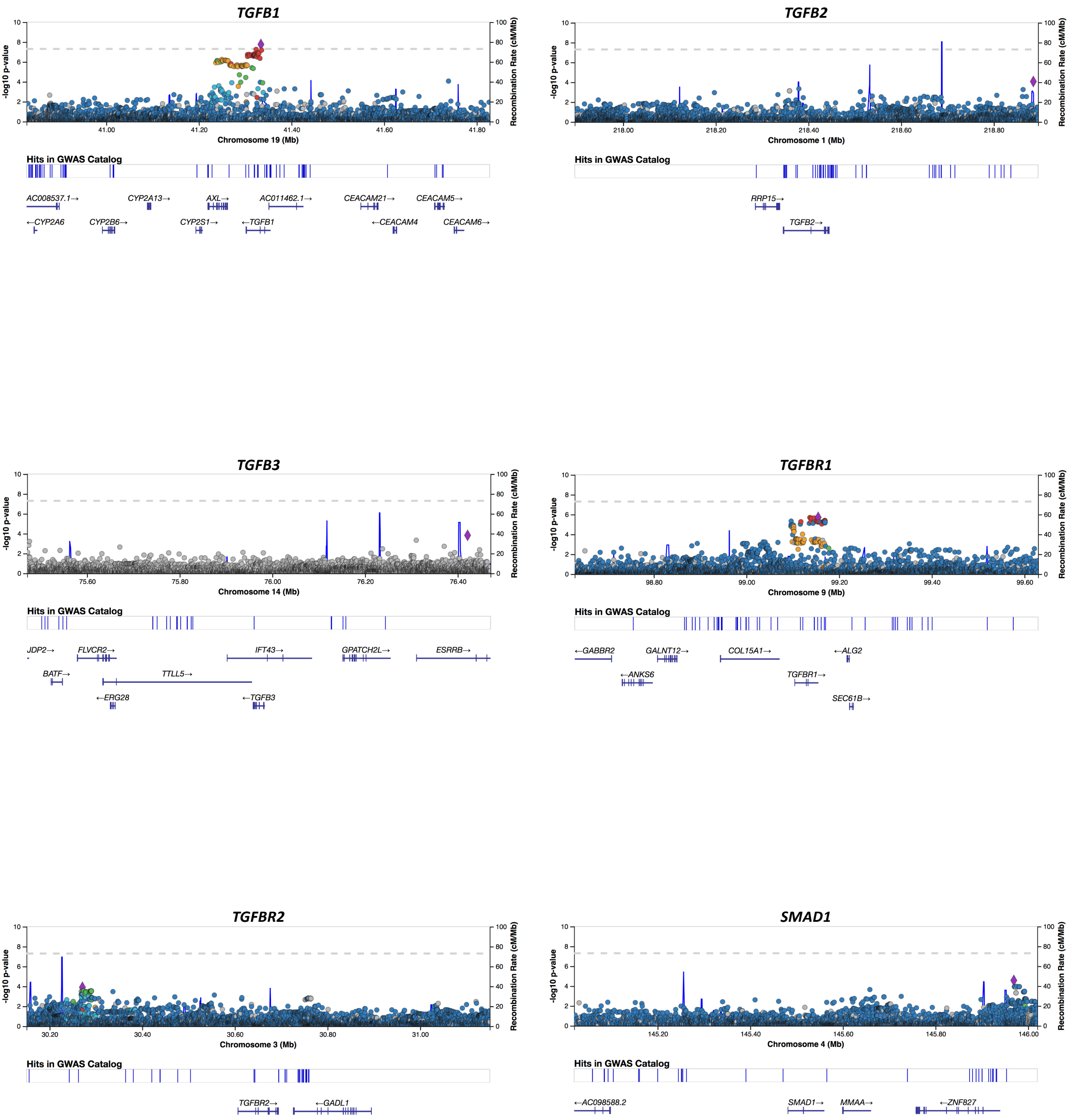

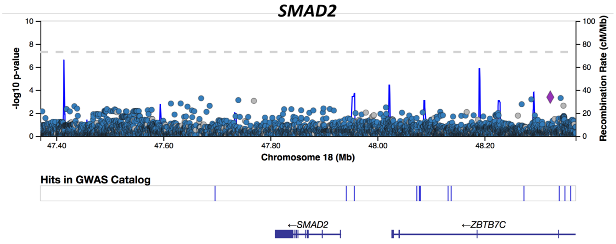

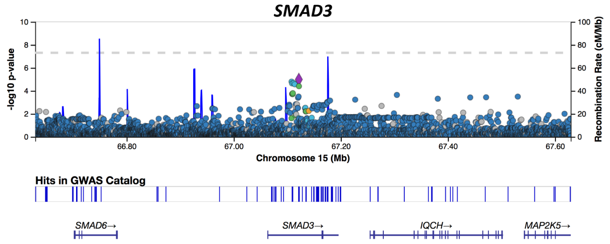

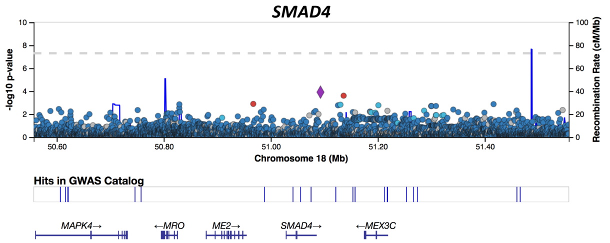

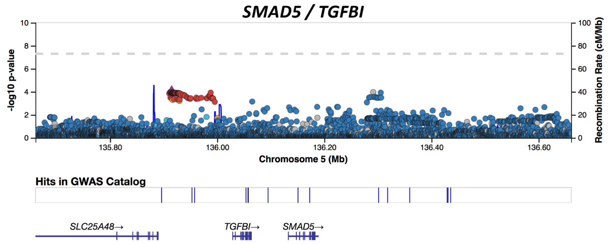

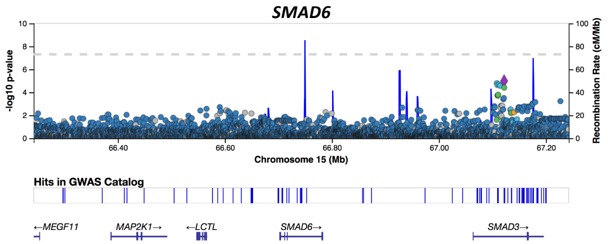

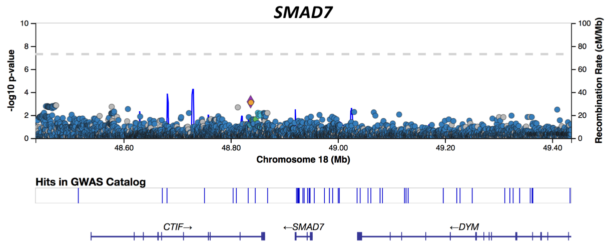

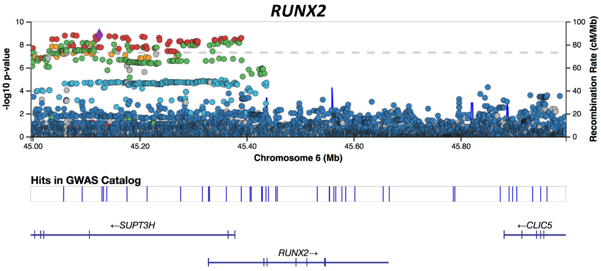

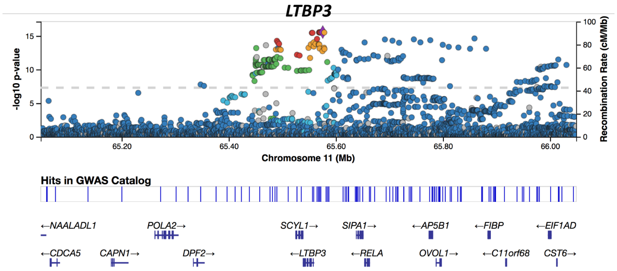

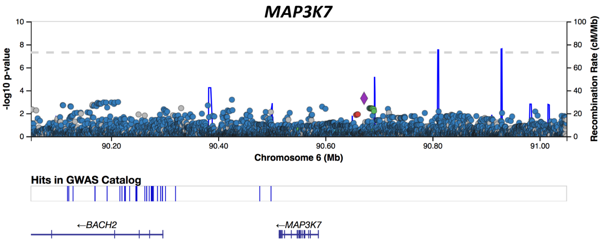

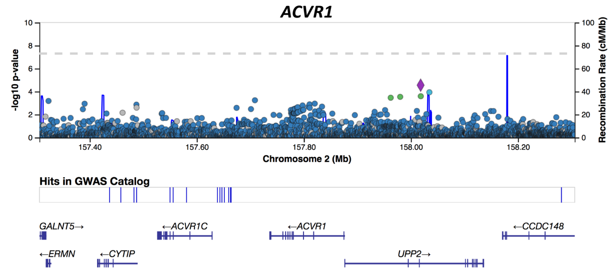

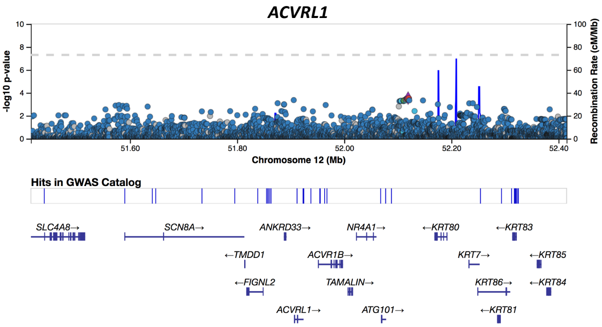

### Supplementary Data 9

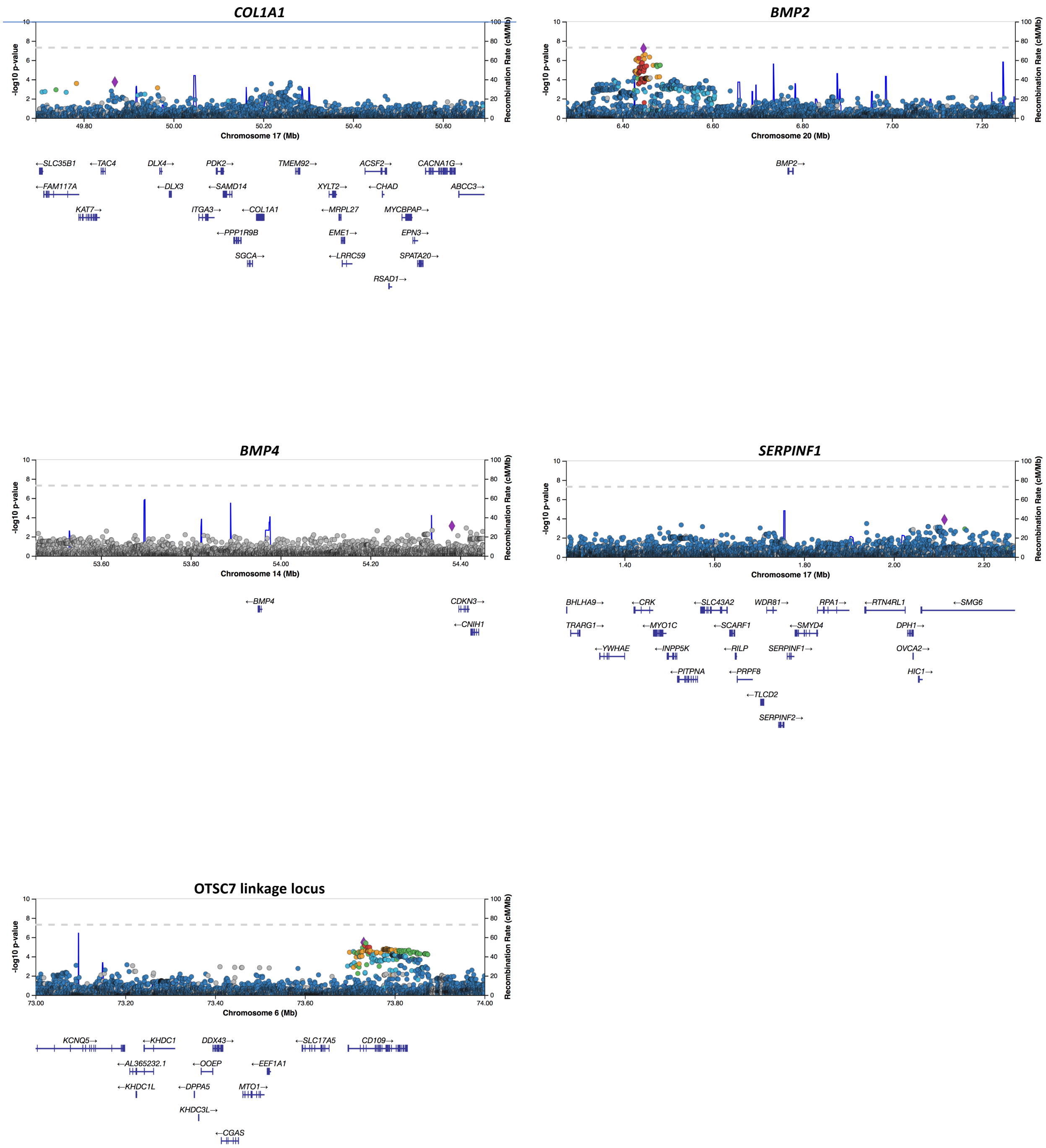
