## Supplementary Data 15 for "Genome-wide Screen of Otosclerosis in Population Biobanks: 18 Loci and Shared Heritability with Skeletal Structure"

**Table 3 References**

1	Wu, M., Chen, G. & Li, Y.-P. TGF-β and BMP signaling in osteoblast, skeletal development, and bone formation, homeostasis and disease. *Bone Research* **4**, 16009, doi:10.1038/boneres.2016.9 (2016).

2	Crane, J. L. & Cao, X. Bone marrow mesenchymal stem cells and TGF-β signaling in bone remodeling. *The Journal of clinical investigation* **124**, 466-472, doi:10.1172/JCI70050 (2014).

3	Kinoshita, A. *et al.* Domain-specific mutations in TGFB1 result in Camurati-Engelmann disease. *Nat. Genet.* **26**, 19-20, doi:10.1038/79128 (2000).

4	Komori, T. Regulation of Proliferation, Differentiation and Functions of Osteoblasts by Runx2. *Int. J. Mol. Sci.* **20**, doi:10.3390/ijms20071694 (2019).

5	Barutcu, A. R. *et al.* The bone-specific Runx2-P1 promoter displays conserved three-dimensional chromatin structure with the syntenic Supt3h promoter. *Nucleic Acids Res.* **42**, 10360-10372, doi:10.1093/nar/gku712 (2014).

6	Moffatt, P. *et al.* Metaphyseal dysplasia with maxillary hypoplasia and brachydactyly is caused by a duplication in RUNX2. *Am. J. Hum. Genet.* **92**, 252-258, doi:10.1016/j.ajhg.2012.12.001 (2013).

7	Komori, T. *et al.* Targeted disruption of Cbfa1 results in a complete lack of bone formation owing to maturational arrest of osteoblasts. *Cell* **89**, 755-764, doi:10.1016/s0092-8674(00)80258-5 (1997).

8	Moffatt, P. *et al.* Metaphyseal dysplasia with maxillary hypoplasia and brachydactyly is caused by a duplication in RUNX2. *Am. J. Hum. Genet.* **92**, 252-258, doi:10.1016/j.ajhg.2012.12.001 (2013).

9	Chang, J. L. *et al.* Tissue-specific calibration of extracellular matrix material properties by transforming growth factor-β and Runx2 in bone is required for hearing. *EMBO Rep* **11**, 765-771, doi:10.1038/embor.2010.135 (2010).

10	Demetriou, M., Binkert, C., Sukhu, B., Tenenbaum, H. C. & Dennis, J. W. Fetuin/alpha2-HS glycoprotein is a transforming growth factor-beta type II receptor mimic and cytokine antagonist. *J. Biol. Chem.* **271**, 12755-12761, doi:10.1074/jbc.271.22.12755 (1996).

11	Szweras, M. *et al.* alpha 2-HS glycoprotein/fetuin, a transforming growth factor-beta/bone morphogenetic protein antagonist, regulates postnatal bone growth and remodeling. *J. Biol. Chem.* **277**, 19991-19997, doi:10.1074/jbc.M112234200 (2002).

12	Schinke, T. *et al.* The serum protein alpha2-HS glycoprotein/fetuin inhibits apatite formation in vitro and in mineralizing calvaria cells. A possible role in mineralization and calcium homeostasis. *J. Biol. Chem.* **271**, 20789-20796, doi:10.1074/jbc.271.34.20789 (1996).

13	Heiss, A. *et al.* Structural basis of calcification inhibition by alpha 2-HS glycoprotein/fetuin-A. Formation of colloidal calciprotein particles. *J. Biol. Chem.* **278**, 13333-13341, doi:10.1074/jbc.M210868200 (2003).

14	Price, P. A. & Lim, J. E. The inhibition of calcium phosphate precipitation by fetuin is accompanied by the formation of a fetuin-mineral complex. *J. Biol. Chem.* **278**, 22144-22152, doi:10.1074/jbc.M300744200 (2003).

15	Rittenberg, B. *et al.* Regulation of BMP-induced ectopic bone formation by Ahsg. *J. Orthop. Res.* **23**, 653-662, doi:10.1016/j.orthres.2004.11.010 (2005).

16	Robertson, I. B. *et al.* Latent TGF-β-binding proteins. *Matrix Biol.* **47**, 44-53, doi:10.1016/j.matbio.2015.05.005 (2015).

17	McInerney-Leo, A. M. *et al.* Mutations in LTBP3 cause acromicric dysplasia and geleophysic dysplasia. *J. Med. Genet.* **53**, 457-464, doi:10.1136/jmedgenet-2015-103647 (2016).

18	Dugan, S. L. *et al.* New recessive truncating mutation in LTBP3 in a family with oligodontia, short stature, and mitral valve prolapse. *Am. J. Med. Genet. A* **167**, 1396-1399, doi:10.1002/ajmg.a.37049 (2015).

19	McInerney-Leo, A. M. *et al.* Mutations in <em>LTBP3</em> cause acromicric dysplasia and geleophysic dysplasia. *J. Med. Genet.* **53**, 457-464, doi:10.1136/jmedgenet-2015-103647 (2016).

20	Huckert, M. *et al.* Mutations in the latent TGF-beta binding protein 3 (LTBP3) gene cause brachyolmia with amelogenesis imperfecta. *Hum. Mol. Genet.* **24**, 3038-3049, doi:10.1093/hmg/ddv053 (2015).

21	Dabovic, B. *et al.* Osteopetrosis-like phenotype in latent TGF-beta binding protein 3 deficient mice. *Bone* **37**, 25-31, doi:10.1016/j.bone.2005.02.021 (2005).

22	Kokura, K. *et al.* The Ski-binding Protein C184M Negatively Regulates Tumor Growth Factor-β Signaling by Sequestering the Smad Proteins in the Cytoplasm. *J. Biol. Chem.* **278**, 20133-20139, doi:10.1074/jbc.M210855200 (2003).

23	Gowen, L. C. *et al.* Targeted disruption of the osteoblast/osteocyte factor 45 gene (OF45) results in increased bone formation and bone mass. *J. Biol. Chem.* **278**, 1998-2007, doi:10.1074/jbc.M203250200 (2003).

24	Gullard, A. *et al.* MEPE Localization in the Craniofacial Complex and Function in Tooth Dentin Formation. *J. Histochem. Cytochem.* **64**, 224-236, doi:10.1369/0022155416635569 (2016).

25	Staines, K. A. *et al.* MEPE is a novel regulator of growth plate cartilage mineralization. *Bone* **51**, 418-430, doi:10.1016/j.bone.2012.06.022 (2012).

26	Schrauwen, I. *et al.* Variants affecting diverse domains of MEPE are associated with two distinct bone disorders, a craniofacial bone defect and otosclerosis. *Genet. Med.* **21**, 1199-1208, doi:10.1038/s41436-018-0300-5 (2019).

27	Chen, X., Zhang, K., Hock, J., Wang, C. & Yu, X. Enhanced but hypofunctional osteoclastogenesis in an autosomal dominant osteopetrosis type II case carrying a c.1856C>T mutation in CLCN7. *Bone Research* **4**, 16035, doi:10.1038/boneres.2016.35 (2016).

28	Waguespack, S. G. *et al.* Chloride channel 7 (ClCN7) gene mutations and autosomal dominant osteopetrosis, type II. *J. Bone Miner. Res.* **18**, 1513-1518, doi:10.1359/jbmr.2003.18.8.1513 (2003).

29	Pangrazio, A. *et al.* Molecular and clinical heterogeneity in CLCN7-dependent osteopetrosis: report of 20 novel mutations. *Hum. Mutat.* **31**, E1071-1080, doi:10.1002/humu.21167 (2010).

30	Chang, E. J. *et al.* Brain-type creatine kinase has a crucial role in osteoclast-mediated bone resorption. *Nat. Med.* **14**, 966-972, doi:10.1038/nm.1860 (2008).

31	Calabrese, G. M. *et al.* Integrating GWAS and Co-expression Network Data Identifies Bone Mineral Density Genes SPTBN1 and MARK3 and an Osteoblast Functional Module. *Cell Systems* **4**, 46-59.e44, doi:<https://doi.org/10.1016/j.cels.2016.10.014> (2017).

32	Matsuoka, K. *et al.* WAIF1 Is a Cell-Surface CTHRC1 Binding Protein Coupling Bone Resorption and Formation. *J. Bone Miner. Res.* **33**, 1500-1512, doi:10.1002/jbmr.3436 (2018).

33	Miyauchi, A. *et al.* Recognition of osteopontin and related peptides by an alpha v beta 3 integrin stimulates immediate cell signals in osteoclasts. *J. Biol. Chem.* **266**, 20369-20374 (1991).

34	Kruger, T. E., Miller, A. H., Godwin, A. K. & Wang, J. Bone sialoprotein and osteopontin in bone metastasis of osteotropic cancers. *Crit. Rev. Oncol. Hematol.* **89**, 330-341, doi:10.1016/j.critrevonc.2013.08.013 (2014).

35	Faccio, R. *et al.* Activation of alphav beta3 integrin on human osteoclast-like cells stimulates adhesion and migration in response to osteopontin. *Biochem. Biophys. Res. Commun.* **249**, 522-525, doi:10.1006/bbrc.1998.9180 (1998).

36	Xiao, Z. *et al.* Osteoblast-specific deletion of Pkd2 leads to low-turnover osteopenia and reduced bone marrow adiposity. *PLoS One* **9**, e114198-e114198, doi:10.1371/journal.pone.0114198 (2014).

37	Ji, S., Wang, S., Zhao, X. & Lv, L. Long noncoding RNA NEAT1 regulates the development of osteosarcoma through sponging miR-34a-5p to mediate HOXA13 expression as a competitive endogenous RNA. *Mol Genet Genomic Med* **7**, e673-e673, doi:10.1002/mgg3.673 (2019).

38	Zhang, Y. *et al.* lncRNA Neat1 Stimulates Osteoclastogenesis Via Sponging miR-7. *J. Bone Miner. Res.* **n/a**, doi:10.1002/jbmr.4039.

39	Tohmonda, T. *et al.* IRE1α/XBP1-mediated branch of the unfolded protein response regulates osteoclastogenesis. *J. Clin. Invest.* **125**, 3269-3279, doi:10.1172/jci76765 (2015).

40	Nakajima, M. *et al.* A genome-wide association study identifies susceptibility loci for ossification of the posterior longitudinal ligament of the spine. *Nat. Genet.* **46**, 1012-1016, doi:10.1038/ng.3045 (2014).

41	Lüdecke, H. J. *et al.* Genotypic and phenotypic spectrum in tricho-rhino-phalangeal syndrome types I and III. *Am. J. Hum. Genet.* **68**, 81-91, doi:10.1086/316926 (2001).

42	Zehnder, A. F., Kristiansen, A. G., Adams, J. C., Merchant, S. N. & McKenna, M. J. Osteoprotegerin in the inner ear may inhibit bone remodeling in the otic capsule. *Laryngoscope* **115**, 172-177, doi:10.1097/01.mlg.0000150702.28451.35 (2005).

43	Wright, H. L., McCarthy, H. S., Middleton, J. & Marshall, M. J. RANK, RANKL and osteoprotegerin in bone biology and disease. *Curr. Rev. Musculoskelet. Med.* **2**, 56-64, doi:10.1007/s12178-009-9046-7 (2009).

44	Zehnder, A. F. *et al.* Osteoprotegrin knockout mice demonstrate abnormal remodeling of the otic capsule and progressive hearing loss. *Laryngoscope* **116**, 201-206, doi:10.1097/01.mlg.0000191466.09210.9a (2006).

45	Priyadarshi, S. *et al.* Genetic Association and Altered Gene Expression of Osteoprotegerin in Otosclerosis Patients. *Ann. Hum. Genet.* **79**, 225-237, doi:10.1111/ahg.12118 (2015).
